# Congregate Shelter Characteristics and Prevalence of Asymptomatic SARS-CoV-2

**DOI:** 10.1101/2020.05.21.20108985

**Authors:** Elizabeth A. Samuels, Rebecca Karb, Rahul Vanjani, M. Catherine Trimbur, Anthony Napoli

**Author notes:** equivalent first author (Elizabeth A. Samuels, Rebecca Karb).

## Abstract

**Background:** Individuals experiencing homelessness residing in congregate shelters have increased risk for SARS-CoV-2 infection. Prevalence of asymptomatic SARS-CoV-2 in congregate shelters is high, but shelter characteristics associated with SARS-CoV-2 transmission are currently unknown.

**Methods:** We conducted a cross-sectional, multicenter cohort study across five congregate shelters in Rhode Island. We tested people 18 years of age and older staying in Rhode Island congregate shelters in April 2020 during the COVID-19 pandemic. A survey instrument was designed and implemented based on an a priori sample size. All consented participants reported basic demographics, recent travel, duration of time at the shelter, any symptomatology, and had their temperature and pulse oximetry measured. Each participant was tested for COVID-19 using nasopharyngeal swabbing. Shelter characteristics about location, occupancy, resident length of stay, and COVID-19 mitigation strategies were collected through structured phone questionnaire with shelter staff.

**Results:** A total of 302 individuals were screened and 299 participated across five homeless shelters. The median age was 47.9 (range 18-85) and 20% were female. Of the 299 participants, 35 (11.7%) were positive for SARS-CoV-2; rates varied among shelters, ranging from 0% to 35%. Among the participants in the study, 5% had a new cough, 4% shortness of breath, and 3% reported loss of taste or smell. Symptom prevalence did not vary significantly between positive and negative SARS-CoV-2 groups. Regular symptom screening was not associated with lower infection rates. Shelters with higher rates of positivity were in more densely populated areas, cared for a more transient populations, and instituted fewer social distancing practices for sleeping arrangements or mealtimes.

**Conclusions and Relevance:** Residents of congregate shelters are at increased risk for asymptomatic transmission of SARS-CoV-2. To reduce transmission and enable continuation of low-threshold shelter services, there is a need for universal testing, implementation of infection control and physical distancing measures within congregate shelters, and expansion of non-congregate supportive housing.

## Background

There are high rates of asymptomatic SARS-CoV-2 in congregate homeless shelters^1,2^, but little is known about shelter-level risk factors and successful mitigation strategies. While many shelters have worked to implement Centers for Disease Control and Prevention (CDC) recommendations to control transmission (e.g. performing daily symptom screening, allowing people to stay at the shelter during the day, and providing on-site meals),^4^ these mitigation strategies can be difficult to implement and have unclear benefits. Given the high estimated rate of asymptomatic and pre-symptomatic transmission^1,4^ coupled with the inherent risks of congregate living, symptom screening alone is insufficient to control the spread of SARS-CoV-2. Comprehensive testing accompanied by safe, stable housing is necessary to mitigate risks for people experiencing homelessness.

## Objective

Describe the varying prevalence of asymptomatic SARS-CoV-2 infection in congregate shelters and associated shelter characteristics and practices.

## Methods and Findings

We conducted a cross-sectional cohort study of congregate shelter residents 18 years of age and older staying in five shelters in the Providence Metro Region, from April 19-April 24, 2020. Testing occurred during the peak of new case identification.^5^ During this time, congregate shelters remained open and people experiencing homelessness positive for SARS-CoV-2 were isolated in a hotel, arranged by the Rhode Island Department of Health (RIDOH) and supported by on-site case management. Shelter characteristics and infection control strategies were assessed by structured telephone interview. Shelter residents consenting for testing were screened for COVID-19 symptoms, concurrent comorbidities, and had temporal temperature and pulse oximetry measured. Data were collected and managed using REDCap (Vanderbilt, Nashville, TN). Nasopharyngeal swabbing was done by an emergency physician with training in appropriate nasopharyngeal swab technique. Tests were run on one of three available assays, Roche (Basel, Switzerland), Cepheid (Sunnyvale, CA), and Abbott (Chicago, IL). The study was deemed exempt by the RIDOH Institutional Review Board.

We used descriptive statistics to summarize participant and shelter characteristics. We compared the proportion of positive SARS-CoV-2 tests among shelters, demographic groups, medical comorbidities, and symptomatology using t-tests and Fisher exact tests using STATA (Statacrop, College Station, TX).

299 individuals were tested for SARS-CoV-2 at five shelters. There were no significant differences in prevalence based on race or gender (Table 1). Shelters 2 and 5 had positivity rates of 35.3% and 21.6%, respectively, while all other shelters had zero confirmed cases (Table 1). There were no statistical differences in symptoms between positive and negative groups (20% vs 14%, p=0.34, Table 2). Only 20% of the confirmed positive cases reported any symptoms, none had a documented fever at time of testing, and there were no differences in oxygen saturation (p=0.59, Table 2).

**Table 1:** Demographics, Shelter Characteristics and Practices, and SARS-CoV-2 results by Congregate Shelter in Greater Providence, Rhode Island.

|  | No (%) |  |  |  |  |  |  |
| --- | --- | --- | --- | --- | --- | --- | --- |
|  |  | Shelter |  |  |  |  |  |
|  | All | 1 | 2 | 3 | 4 | 5 | P-value |
| Number Tested | 299 | 48 | 68 | 89 | 43 | 51 |  |
| SARS-CoV-2 + | 35 (11.7) | 0 | 24 (35.3) | 0 | 0 | 11 (21.6) | 0.000* |
| **<u>Shelter Demographics</u>** |  |  |  |  |  |  |  |
| Age, mean (range) | 47.9<br>(18-85) | 47.8<br>(25-76) | 46.7<br>(19-69) | 48.5<br>(20-72) | 53.7<br>(30-85) | 43.4<br>(18-67) | 0.13 |
| Female | 59 (20) | 32 (67) | 9 (13) | 0 | 0 | 18 (35) | 0.000* |
| Race |  |  |  |  |  |  |  |
| Black | 59 (20) | 5 (10) | 17 (25) | 17 (19) | 9 (21) | 11 (22) | 0.01* |
| White | 160 (53) | 35 (73) | 36 (53) | 50 (56) | 21 (49) | 18 (36) |  |
| AI/ Alaska Native | 10 (3) | 3 (6) | 3 (4) | 3 (3) | 0 | 1 (2) |  |
| Other/Unknown | 70 (23) | 5 (10) | 12 (18) | 19 (21) | 13 (30) | 21 (40) |  |
| Latino/a/x |  |  |  |  |  |  |  |
| Latino/a/x | 68 (23) | 11 (23) | 12 (18) | 15 (17) | 16 (37) | 14 (27) | 0.01* |
| Non Latino/a/x | 213 (71) | 37 (77) | 54 (79) | 66 (74) | 27 (63) | 29 (57) |  |
| Other/Unknown | 18 (6) | 0 | 2 (3) | 8 (9) | 0 | 8 (15) |  |
| % Reporting medical comorbidities | 38 | 63 | 34 | 42 | 37 | 13 | 0.000* |
| % Reporting symptoms | 14.7 | 19 | 28 | 8 | 12 | 8 | 0.01* |
| % positive reporting symptoms | 20 | N/A | 30 | N/A | N/A | 0 | 0.000* |
| % Fever (T>100.4) | 0 | 0 | 0 | 0 | 0 | Unknown<sup>a</sup> | — |
| % Hypoxia (O2 sat<92%) | 8 | 6 | 1 | 1 | 0 | Unknown<sup>a</sup> | 0.001* |
| **<u>Shelter Characteristics</u>** |  |  |  |  |  |  |  |
| Census tract pop density<br>(per/sq mile) |  | 10,852 | 21,645 | 2,753 | 2,362 | 10,852 |  |
| % Census Tract multi-unit housing |  | 89 | 90 | 21 | 69 | 89 |  |
| % Of bed capacity (previous night) |  | 90 | 97 | 88 | 100 | 100 |  |
| % At shelter >14d |  | 96 | 58 | 82 | 98 | Unknown<sup>a</sup> |  |
| **<u>Preventative Measures</u>** |  |  |  |  |  |  |  |
| Staff and residents wear masks |  | Yes | Yes | Yes | Yes | Yes |  |
| Daily temperature checks |  | Yes | Yes | Yes | Yes | Yes |  |
| Symptoms screenings |  | Daily | Daily | 2x/daily | 2x/daily | Daily |  |
| Onsite meals offered |  | Yes | Yes | Yes | Yes | Yes |  |
| Social distancing with sleeping spaces |  | Yes | No | No | Yes | Yes |  |
| Open 24 hours |  | Yes | Yes | Yes | Yes | Yes |  |
| Daily education/updates |  | No | No | Yes | No | No |  |
| New residents allowed |  | Yes | Yes | No | No | Yes |  |

<sup>a</sup> Testing was done at this shelter just prior to addition of this data

**Table 2:**
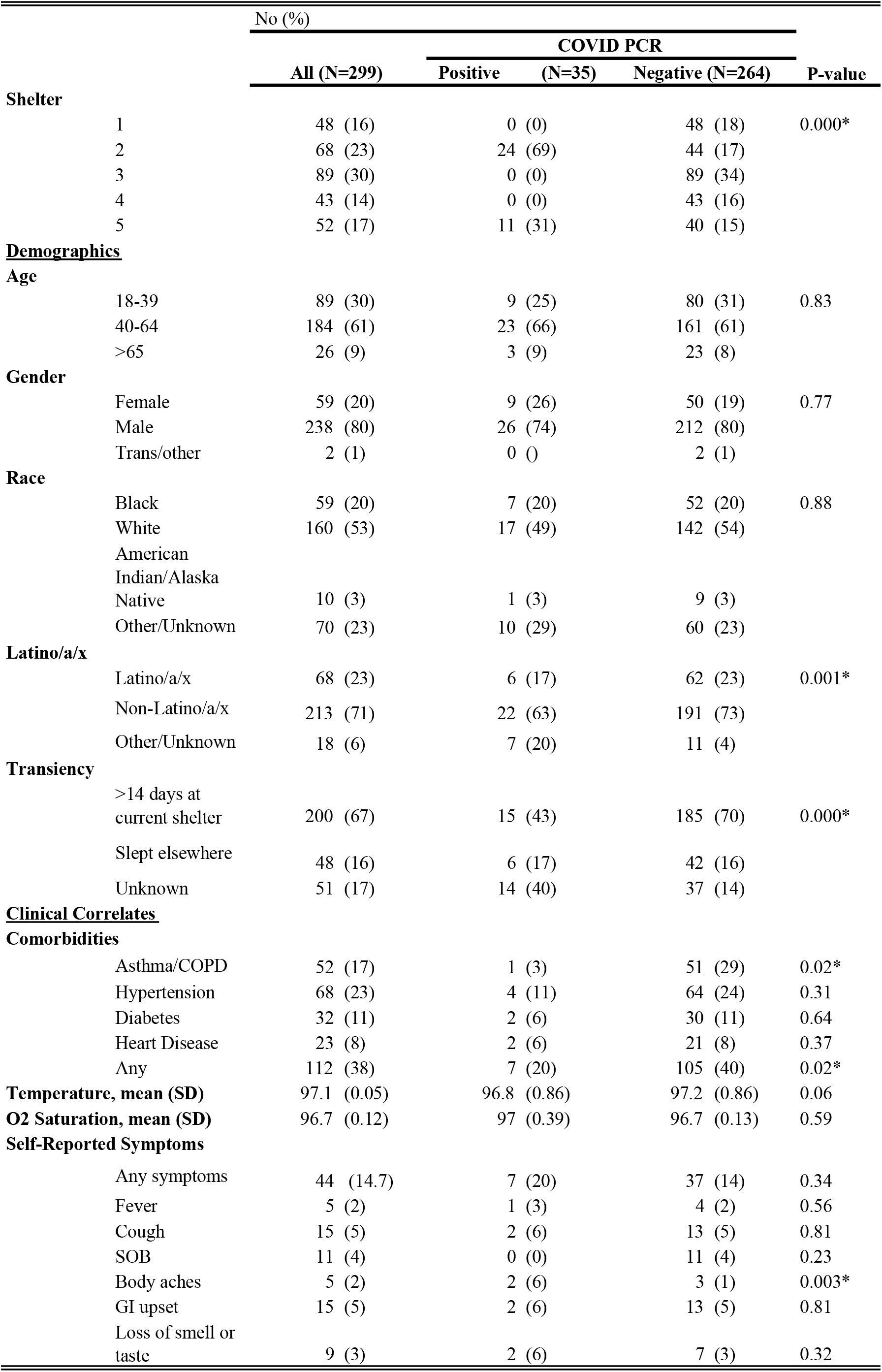
Demographics and Clinical Characteristics of Shelter Residents by SARS-CoV-2 result.

The shelters with positive cases have several distinct characteristics (Table 1). These shelters’ neighborhoods had higher census tract population densities, the shelters were at almost full capacity, and they allowed new residents. Furthermore, Shelter 2’s resident population was more transient, with only 58% reporting staying there for more than 2 weeks.

## Discussion

Asymptomatic carriage of SARS-CoV-2 in congregate shelters does not impact all shelters equally. Our study is limited by a small number of shelters; however, several interesting observations emerged. Shelters with more transient residents had higher prevalence rates. Shelters in locations with lower population density and who limited new residents during the outbreak had zero prevalence in our sample. At the time of our study, many shelter residents who had tested positive were already housed in a hotel, which likely led to an underestimate of true prevalence of COVID in the unhoused population. Furthermore, testing done at the shelters with more transient residents only reflects the residents staying there on the night of testing, not the entire group that intermittently utilizes shelter services.

The range of asymptomatic prevalence of COVID-19 found in different shelters builds on growing data from other cities and has important policy implications. Shelters initially relied on symptom screening to control spread of SARS-CoV-2. Our results add to growing evidence that symptom screening and temperature monitoring are insufficient means to mitigate transmission of SARS-CoV-2 in congregate settings. Sheltering in place, wearing masks, and physical distancing may be at least partially effective in shelters with lower occupancy and a more stable resident population. However, low-threshold shelters provide an important safety net to many people experiencing homelessness and shelters cannot be closed without readily available supportive housing. In order to prevent SARS-CoV-2 transmission while continuing to provide essential, accessible services to people experiencing homelessness, there is a need for universal testing, infection control support at congregate shelters, and expanded availability of safe and supportive housing.

## Data Availability

Data available upon request.

## Acknowledgements

For their collaboration and assistance, the authors would like to acknowledge and thank the Rhode Island Health State Laboratory, the Lifespan Clinical Microbiology Lab, Dr. Utpala Bandy, Dr. Philip Chan, Dr. Kim Chapin, Amy Ferguson, Deborah Garneau, MA, Sari Greene, Dr. Ewa King, Dr. Louis Marchetti, Marilyn McAllister, and Dr. James McDonald. For their assistance with test conduction and data collection, we thank Dr. Jay Baruch, Dr. Michelle Breda, Natalie Chadwick, PA, Dr. Hannah Chason, Dr. Rachel Cooper, Dr. Catherine Cummings, Dr. Giovanna Deluca, Dr. Elizabeth Goldberg, Dr. Meredith Horton, Dr. Ilse Jenouri, Dr. Josh Kaine, Dr. Anita Knopov, Dr. Larry Kogan, Dr. Victoria Leytin, Dr. Tracy Madsen, Dr. Christina Matulis, Dr. Caroline Meehan, Dr. Rory Merritt, Dr. Luke Messac, Dr. Michelle Myles, Dr. Elizabeth Nestor, Dr. David Portelli, Amy Riendeau, NP, Dr. Mahawa Sam, Dr. Jeremiah Schuur, Dr. Lynn Sweeney, and Dr. Michael Tcheyan.

## References

1. Baggett TP, Keyes H, Sporn N, et al. Prevalence of SARS-CoV-2 Infection in Residents of a Large Homeless Shelter in Boston. JAMA. 2020.

2. Mosites E, Parker EM, Clarke KEN, et al. Assessment of SARS-CoV-2 Infection Prevalence in Homeless Shelters — Four U.S. Cities, March 27–April 15, 2020. MMWR Morbidity and Mortality Weekly Report. 2020;69(17).

3. National Center for Immunization and Respiratory Diseases (NCIRD) DoVD. Interim Guidance for Homeless Service Providers to Plan and Respond to Coronavirus Disease 2019 (COVID-19). Centers for Disease Control and Prevention,. https://www.cdc.gov/coronavirus/2019-ncov/community/homeless-shelters/plan-prepare-respond.html. Published 2020. Accessed Aptil 26, 2020.

4. Bai Y, Yao L, Wei T, et al. Presumed Asymptomatic Carrier Transmission of COVID-19. Jama. 2020.

5. Rhode Island Department of Health Center for Health Data and Analysis. Rhode Island COVID-19 Response Data https://ri-department-of-health-covid-19-data-rihealth.hub.arcgis.com. Published 2020. Accessed April 26, 2020.

